# Surrogate Endpoint Evaluation with Causal Mediation: Lessons from the A4 Trial

**DOI:** 10.64898/2026.07.23.26358810

**Authors:** Evan J. Hoefen, Michael D. Flanders, Jason R. Gantenberg, Eleanor Hayes-Larson, Paul K. Crane, Seo-Eun Choi, Emily H. Trittschuh, Sarah F. Ackley

## Abstract

Surrogate endpoints, or measures used in place of the true outcome of interest, have relevance across multiple disease areas. The Prentice Criteria, proposed in 1989, assess surrogacy by evaluating how the treatment’s effect on the true outcome operates through the potential surrogate. Using the A4 Study of solanezumab, we evaluate multiple formulations of the Prentice Criteria using causal mediation. We estimated direct and indirect effects of solanezumab on cognitive decline through cerebral amyloid across different, but reasonable, measures of exposure, mediator, outcome, and covariate adjustment. Unsurprisingly given that solanezumab did not show benefit, estimated indirect effects were close to zero. These results provide little evidence of meaningful mediation for memory and global cognition, but precision varied substantially. Confidence interval widths varied by up to a factor of 17 across specifications. Causal mediation analysis of individual-level randomized trial data may contribute to quantitative surrogate endpoint evaluation, but our findings indicate this is only the case when analytic choices are biologically justified, prespecified, and interpreted with attention to uncertainty.

## Introduction

Surrogate endpoints, or measures used in place of true outcomes of interest because they can be measured earlier, have broad relevance across disease areas. In 1989, Prentice proposed criteria for evaluating whether a candidate surrogate could substitute for a true outcome in testing treatment effects (Prentice, 1989). These criteria remain widely cited because they formalize the concept that a valid surrogate should not merely be associated with the outcome, but should mediate the treatment–outcome relationship. The Prentice “capture” criterion is often read as full mediation: treatment affects the surrogate, the surrogate affects the outcome, and treatment has no residual effect after accounting for the surrogate. However, the Prentice criteria predated modern mediation methods. Causal mediation analysis can therefore operationalize “capture.” FDA decisions about surrogate endpoints do not depend on formal demonstration that the Prentice criteria are met, in part because the criteria are di”cult to satisfy and operationalize in practice (Berger, 2004; Heller, 2015; Fleming and DeMets, 1996).

Instead, the FDA relies on case-specific judgment informed by biological plausibility, disease context, and prior evidence (Pease et al., 2017).

However, historical surrogate endpoint failures underscore the need for rigorous strategies for evaluation (e.g.,Echt et al., 1991). Alzheimer’s disease is a particularly important setting for this problem. The FDA has approved treatments based on changes in cerebral amyloid, a putative surrogate endpoint, despite significant controversy and calls for more formal surrogate evaluation (Ren et al., 2026; Ackley, M. D. Flanders, et al., 2026; Fairchild and McDaniel, 2017).

We use individual-level data from the A4 Study of solanezumab, currently the only publicly available phase 3 data of a drug that reduces cerebral amyloid (Sperling et al., 2023). We used casual mediation analyses to determine whether amyloid (potential surrogate) mediates the effect of solanezumab on cognitive decline. We evaluate the extent to which mediation-based operationalizations of the Prentice criteria depend on measures used. Here, we attempt to underscore the challenges with implementing the Prentice criteria in practice prior to the release of additional trial data of effective drugs.

## Methods

### Cohort

We used data from the Anti-Amyloid Treatment in Asymptomatic Alzheimer’s (A4) tiral. Participants were randomized to receive either solanezumab or placebo (Sperling et al., 2023). Study enrollment began in 2014 and 1169 participants aged 65-86 were randomized. A4 study data were obtained from the A4/LEARN Study Data Package (A4studydata.org).

### Exposures

The primary exposure was randomization to treatment. In addition to this, we used a variable that captured variation in cumulative treatment exposure arising from a protocol-approved dose escalation. In June 2017, an increase in the study drug from 400 mg to 1600 mg was approved. Thus, depending on when a participant in the treated group enrolled, they can be considered pseudorandomized to a total dose of the drug that depended on the time from individual randomization to treatment escalation.

### Mediators

Mediators evaluated were change in cerebral amyloid evaluated using positron emission tomography (amyloid-PET), change in peripheral phosphorylated tau 217 (p-tau 217), and follow-up p-tau 217. P-tau 217 is a blood-based measure and validated measure of cerebral amyloid more correlated with cerebral amyloid than amyloid-based blood measures (Hayes-Larson et al., 2024).

### Outcomes

They study’s primary cognitive outcome was change the Preclinical Alzheimer Cognitive Composite (PACC), a composite measure of global cognition. As our second cognitive outcome we use change in memory scores obtained from the Center for Psychometric Analyses in Aging and Neurodegeneration (CPAAN) at the University of Washington. CPAAN generates co-calibrated cognitive scores using modern psychometric approaches. (Mukherjee et al., 2023; Hampton et al., 2023). All cognitive were used to calculate a linear rate of cognitive change for each individual (see Ackley, M. Flanders, et al., 2025). See supplement for broader discussion of change scores and baseline adjustment.

### Analysis

Complete followup data for all analytic variables was available for 830 participants. We used casual mediation analyses to determine whether amyloid (potential surrogate) mediates the effect of solanezumab on cognitive decline (VanderWeele, 2016) by estimating the direct and indirect effects of randomization to solanezumab on rate of cognitive change through amyloid measures. We report natural indirect effects and natural direct effects. Analyses were done in R using the causal mediation package (Tingley et al., 2014).

We did this using an illustrative set of combinations of different versions of the treatment variable, amyloid, and cognitive change measures (Figure 1A). Mediation analyses were performed with and without adjustment for covariates age, sex, baseline amyloid burden, and number of *APOE* -ε4 alleles in both linear mediator and linear outcome models. The treatment escalation variable was rescaled such that a one-unit change in that variable corresponded to the same cumulative solanezumab dosage for the group randomized to treatment. Mediator and outcome variables were *z*-standardized by subtracting the sample mean and dividing by the sample standard deviation, allowing direct and indirect effect estimates to be compared on a common standard-deviation scale.

**Figure 1.**
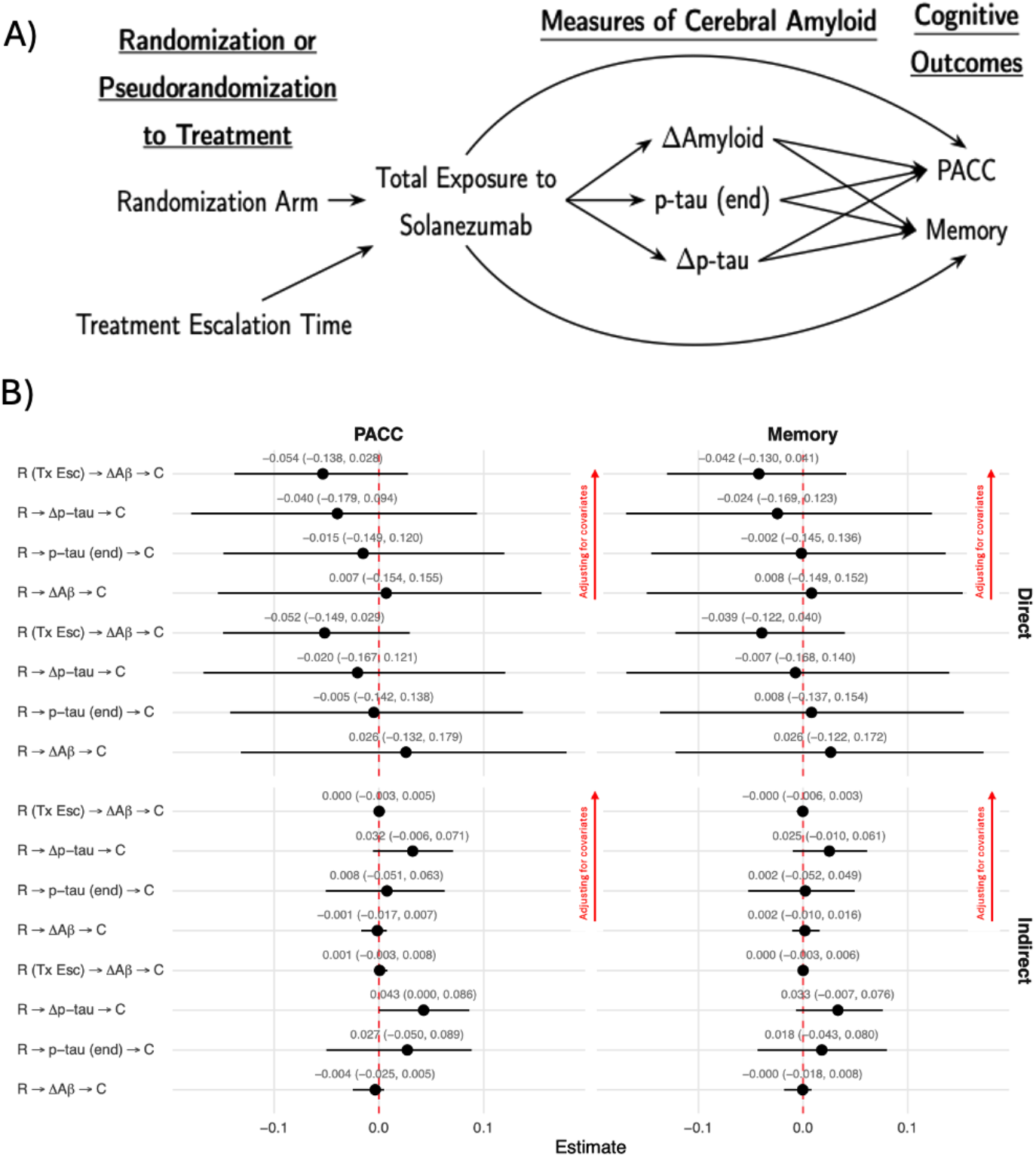
(**a**) Directed acyclic graph used to estimate the extent to which the effect of treatment randomization or pseudo-randomization on cognition operates through amyloid change. The indirect effect represents the component of the effect of randomization on cognition operating through the specified mediator, whereas the direct effect represents the component not operating through that mediator. (**b**) Estimated direct and indirect effects with 95% confidence intervals for two cognitive outcomes: PACC (left) and memory (right). Rows correspond to separate mediation models evaluating amyloid pathways. Points indicate effect estimates and horizontal bars indicate 95% confidence intervals; the dashed vertical line marks the null value. In the model labels, R corresponds to treatment randomization, R (TX esc) corresponds to pseudo-randomization due to treatment escalation, and C denotes the cognitive outcome. Mediators include change in amyloid burden (Δ A *β*), end-of-treatment phosphorylated tau (p-tau end), and change in phosphorylated tau (Δ p-tau).

**Table 1:** Characteristics of individuals in the A4 cohort with follow-up amyloid PET with standard deviatons (SD) or percents (%).

| Characteristic | Placebo | Solanezumab |
| --- | --- | --- |
|  | N = 424 | N = 406 |
|  | N, mean, or % (SD or %) | N, mean, or % (SD or %) |
| N | 424 | 406 |
| Sex (%M) | 40.3% | 41.4% |
| Sex (%F) | 59.7% | 58.6% |
| Age | 71.45 (4.45) | 71.52 (4.47) |
| $\epsilon 4$ allele count | | |
| 0 alleles | 178 (42.0%) | 166 (40.9%) |
| 1 allele | 213 (50.2%) | 203 (50.0%) |
| 2 alleles | 33 (7.8%) | 37 (9.1%) |
| Baseline amyloid (Centiloids) | 58.88 (41.91) | 56.23 (41.26) |
| Baseline p-tau | 0.27 (0.14) | 0.28 (0.16) |
| Baseline PACC | 0.00 (1.00) | 0.00 (1.00) |
| Baseline Memory | 0.42 (0.39) | 0.42 (0.40) |

Since the the A4 trial did not demonstrate overall benefit, percentages mediated were not calculated since estimates can be unstable. However, the decomposition of total effects into direct and indirect effects is still valid and informative as to whether direct and indirect effects are in opposing directions. In a prior publication, we tested for exposure-mediator interactions in our primary model and did not find evidence for interaction (Ackley, M. Flanders, et al., 2025). Confidence intervals were calculated using the bias-corrected and accelerated bootstrap (1000 replicates).

## Results

Estimated standardized direct and indirect effects with 95% confidence intervals (CIs) are given in Figure 1B. Across all models, the estimated indirect effects of solanezumab through amyloid were small, ranging from − 0.004 to 0.043, suggesting little evidence of a substantial mediated effect through amyloid on either outcome.

Precision, however, varied dramatically. The CI widths varied by a factor of 17 between the most and least precisely estimated indirect effects. The highest precision model was pseudorandom treatment escalation *→* change in amyloid as measured with PET *→* memory with covariate adjustment, 95% CI: [− 0.003, 0.005], 95% CI width = 0.008. The lowest precision model was randomization *→* p-tau 217 at the trial end *→* PACC without covariate adjustment, 95% CI: − 0.050, 0.089], 95% CI width = 0.139. Less variability in precision was seen with direct effects than indirect effects. Results from a stability analysis with variable sample sizes based on data availability were qualitatively unchanged (Figure S1).

## Discussion

We performed multiple reasonable versions of mediation analyses to validate amyloid as a surrogate using different measures of randomization to treatment, amyloid, and cognition. Across models, indirect effect estimates were consistently close to zero, providing little evidence that solanezumab affected cognitive decline through amyloid in this trial specifically. However, we found that the precision of those estimates varied dramatically across choices of exposure and mediator.

Causal mediation operationalizes the Prentice criteria using modern methods, but doing so requires choices about exposure, mediator, outcome, and adjustment. Even in the same trial, these choices can substantially alter the precision of the resulting surrogacy assessment. Thus, measure choices changed the evidentiary strength of the surrogate evaluation because a precise near-null estimate supports a different conclusion than an imprecise estimate centered near the same value. This is especially important for regulation, where surrogate acceptability may depend not only on estimated effects but also on uncertainty around those effects (Saraf, Mathew, and Roy, 2015; Buyse et al., 2000). Variability in evidentiary strength could be expected to be more dramatic in studies with an overall effect.

Our findings also help contextualize FDA’s approach to surrogate endpoints (Pease et al., 2017). Regulatory decisions do not require formal fulfillment of the Prentice criteria. This flexibility may reflect the practical di”culty of applying a single highly stringent formal set of surrogacy criteria in a context where different exposure, mediator, outcome, and adjustment choices can be made.

The analytic variability observed here should not be taken as evidence against mediation-based surrogate evaluation, but as evidence that such analyses require prespecified and clinically justified choices of exposure, mediator, outcome, and adjustment set. Principled application of causal mediation analysis to individual-level randomized trial data may yet be a strong component of a robust quantitative surrogate evaluation framework, particularly when paired with transparent prespecification and stability analyses.

Several methodological issues remain unresolved. Existing approaches, including methods to correct for measurement error in the mediator, could help account for observed differences between amyloid measured by p-tau 217 and PET (Ackley, La Joie, et al., 2026; VanderWeele, Valeri, and Ogburn, 2012). However, to the best of our knowledge, existing methods do not simultaneously address differences in exposure definition, mediator measurement, outcome measurement, and adjustment strategy within a unified surrogate endpoint evaluation framework (Burzykowski, Molenberghs, and Buyse, 2005).

This study has limitations. A4 did not demonstrate an overall cognitive benefit of solanezumab, so our analyses should be interpreted as a stress test for mediation-based surrogate evaluation rather than as validation test of amyloid surrogacy. We plan to repeat these analyses in CLARITY-AD and TRAILBLAZER-ALZ 2, trials that showed e”cacy of an amyloid-targeting drug, if and when those data become available. We only performed complete-case analyses, keeping the sample size constant across analyses.

Given the high public health and financial stakes of drug development, quantitative surrogate endpoint evaluation is essential. Invalid surrogates may redirect resources toward interventions patient benefit, while well-validated surrogates could reduce trial duration, lower development costs, and accelerate access to effective therapies (Largent et al., 2022; Fleming and Powers, 2012; Budish, Roin, and Williams, 2015). Our analyses in A4 suggest that causal mediation approach to analysis of individual-level randomized trial data may contribute to this goal, but only when pre-specified and biologically justified analytic choices are made (Ciani et al., 2023).

## Data Availability

A4 data are publicly available to qualified researchers with a short application at a4studydata.org.

https://www.a4studydata.org/

## Funding

This project was supported by the National Institute on Aging grants: R00AG073454, R00AG075317, and P01AG082653.

### AI Disclosure

An original draft of this manuscript was human written by EH and SFA. One co-author subsequently suggested revisions to the draft with text edited using Claude Opus 4.8. The final draft was reviewed and approved by all authors, who take full responsibility for the content of the manuscript.

## Acknowledgments

The A4 Study was a secondary prevention trial in preclinical Alzheimer’s disease, aiming to slow cognitive decline associated with brain amyloid accumulation in clinically normal older individuals. The A4 Study was funded by a public-private-philanthropic partnership, including funding from the National Institutes of Health– National Institute on Aging, Eli Lilly and Company, Alzheimer’s Association, Accelerating Medicines Partnership, GHR Foundation, an anonymous foundation, and additional private donors, with in-kind support from Avid Radiopharmaceuticals, Cogstate, Albert Einstein College of Medicine and the Foundation for Neurologic Diseases. The companion observational Longitudinal Evaluation of Amyloid Risk and Neurodegeneration (LEARN) Study was funded by the Alzheimer’s Association and GHR Foundation. The A4 and LEARN Studies were led by Dr. Reisa Sperling at Brigham and Women’s Hospital, Harvard Medical School, and Dr. Paul Aisen at the Alzheimer’s Therapeutic Research Institute (ATRI) at the University of Southern California. The A4 and LEARN Studies were coordinated by ATRI at the University of Southern California, and the data are made available under the auspices of Alzheimer’s Clinical Trial Consortium through the Global Research & Imaging Platform (GRIP). The complete A4 Study Team list is available on: https://www.actcinfo.org/a4-study-team-lists/. We would like to acknowledge the dedication of the study participants and their study partners who made the A4 and LEARN Studies possible.

## References

Ackley, Sarah F, Michael Flanders, et al. (2025). “Reanalysis of the A4 Study to Formally Evaluate Amyloid Removal as a Surrogate for Cognitive Decline”. In: Alzheimer’s & Dementia 21, e110201.

Ackley, Sarah F, Michael D Flanders, et al. (2026). “Evaluation of amyloid change as a surrogate for cognitive decline: demonstration in individual-level data from the A4 study of solanezumab”. In: Alzheimer’s & Dementia: Translational Research & Clinical Interventions 12.1, e70205.

Ackley, Sarah F, Renaud La Joie, et al. (2026). “Substituting blood-based biomarkers for imaging measures in Alzheimer’s disease studies: implications for sample size and bias”. In: The Journals of Gerontology, Series A: Biological Sciences and Medical Sciences 81.5, glag068.

Berger, Vance W (2004). “Does the Prentice criterion validate surrogate endpoints?” In: Statistics in medicine 23.10, pp. 1571–1578.

Budish, Eric, Benjamin N Roin, and Heidi Williams (2015). “Do firms underinvest in long-term research? Evidence from cancer clinical trials”. In: American Economic Review 105.7, pp. 2044–2085.

Burzykowski, Tomasz, Geert Molenberghs, and Marc Buyse (2005). The evaluation of surrogate endpoints. Springer Science & Business Media.

Buyse, Marc et al. (2000). “Statistical validation of surrogate endpoints: problems and proposals”. In: Drug Information Journal 34.2, pp. 447–454.

Ciani, Oriana et al. (2023). “A framework for the definition and interpretation of the use of surrogate endpoints in interventional trials”. In: EClinicalMedicine 65.

Echt, Debra S et al. (1991). “Mortality and morbidity in patients receiving encainide, flecainide, or placebo: the Cardiac Arrhythmia Suppression Trial”. In: New England journal of medicine 324.12, pp. 781–788.

Fairchild, Amanda J and Heather L McDaniel (2017). “Best (but oft-forgotten) practices: mediation analysis”. In: The American journal of clinical nutrition 105.6, pp. 1259–1271.

Fleming, Thomas R and David L DeMets (1996). “Surrogate end points in clinical trials: are we being misled?” In: Annals of internal medicine 125.7, pp. 605–613.

Fleming, Thomas R and John H Powers (2012). “Biomarkers and surrogate endpoints in clinical trials”. In: Statistics in medicine 31.25, pp. 2973–2984.

Hampton, Olivia L et al. (2023). “Harmonizing the preclinical Alzheimer cognitive composite for multicohort studies.” In: Neuropsychology 37.4, p. 436.

Hayes-Larson, Eleanor et al. (2024). “Considerations for use of blood-based biomarkers in epidemiologic dementia research”. In: American journal of epidemiology 193.3, pp. 527–535.

Heller, G (2015). “Statistical controversies in clinical research: an initial evaluation of a surrogate end point using a single randomized clinical trial and the Prentice criteria”. In: Annals of Oncology 26.10, pp. 2012–2016.

Largent, Emily A et al. (2022). “Aspiring to reasonableness in accelerated approval: anticipating and avoiding the next aducanumab”. In: Drugs & aging 39.6, p. 389.

Mukherjee, Shubhabrata et al. (2023). “Cognitive domain harmonization and cocalibration in studies of older adults.” In: Neuropsychology 37.4, p. 409.

Pease, Alison M et al. (2017). “Postapproval studies of drugs initially approved by the FDA on the basis of limited evidence: systematic review”. In: bmj 357.

Prentice, Ross L (1989). “Surrogate endpoints in clinical trials: definition and operational criteria”. In: Statistics in medicine 8.4, pp. 431–440.

Ren, Sa et al. (2026). “Evaluating amyloid-beta as a surrogate endpoint in trials of anti-amyloid-beta drugs in Alzheimer’s disease: a Bayesian meta-analysis”. In: Journal of Comparative Effectiveness Research 15.1, e250095.

Saraf, Sanatan, Thomas Mathew, and Anindya Roy (2015). “Statistical validation of surrogate endpoints: another look at the Prentice Criterion and Other Criteria”. In: Journal of Biopharmaceutical Statistics 25.6, pp. 1234–1246.

Sperling, Reisa A et al. (2023). “Trial of solanezumab in preclinical Alzheimer’s disease”. In: New England Journal of Medicine 389.12, pp. 1096–1107.

Tingley, Dustin et al. (2014). “mediation: R package for causal mediation analysis”. In: Journal of Statistical Software 59.5, pp. 1–38.

VanderWeele, Tyler J (2016). “Explanation in causal inference: developments in mediation and interaction”. In: International journal of epidemiology 45.6, pp. 1904–1908.

VanderWeele, Tyler J, Linda Valeri, and Elizabeth L Ogburn (2012). “Commentary: the role of measurement error and misclassification in mediation analysis: mediation and measurement error”. In: Epidemiology 23.4, pp. 561–564.

