## Supplemental Information for "Surrogate Endpoint Evaluation with Causal Mediation: Lessons from the A4 Trial"

#### CPAAN

CPAAN is generating co-calibrated scores for memory, executive functioning, language, and visuospatial abilities using modern psychometric approaches as the Cognition Core for the Alzheimer's Disease Sequencing Project Phenotype Harmonization Consortium (ADSP-PHC) (Mukherjee et al., 2023; Hampton et al., 2023). Methods detailing this approach have been previously published and are intended to produce more precise, less noisy measures of cognitive performance, as well as uncertainty quantification.

#### Change Scores

In our mediation analyses, we used change scores as potential mediators (*e.g.*, $\Delta$p-tau, $\Delta$A$\beta$). The *change score* is defined as the value of a given biomarker at follow-up minus its value at baseline. The use of change scores as outcomes has been the subject of spirited debate and disagreement in the epidemiologic literature (Glymour et al., 2005; Tennant et al., 2021; Glymour, 2022). Some authors have argued that modeling the difference in change scores may not correspond to meaningful counterfactual contrasts (Tennant et al., 2021), while others have noted shortcomings in the alternative, statistical adjustment for the baseline value of the outcome (Glymour et al., 2005; Glymour, 2022). These controversies do not pertain solely to observational analyses (Senn, 2006), and pitfalls associated with each approach could conceivably affect our estimates. However, while Glymour et al. 2005 argued that baseline adjustment could induce collider bias when studying an exposure that precedes (and causes) the outcome value at baseline, in a randomized controlled trial baseline amyloid load cannot have been caused by the exposure (in our case, solanezumab). We largely ignore debates regarding change scores, considering our illustrative aims, though we note the similarity in the mediation results when using p-tau at the end of follow-up versus $\Delta$p-tau (Figures 1 and 2).

#### Supplemental Figure

In the primary analyses, we restricted the sample to participants with complete data across all models to maintain a constant analytic sample. This facilitated direct comparison across estimates.

As a stability analysis, we repeated the primary analyses, allowing sample sizes to vary across models. Specifically, we used analytic samples based on follow-up data availability. Each model included all participants with the required baseline, amyloid mediator, and follow-up cognitive data. Results were qualitatively unchanged from primary complete-case analyses.

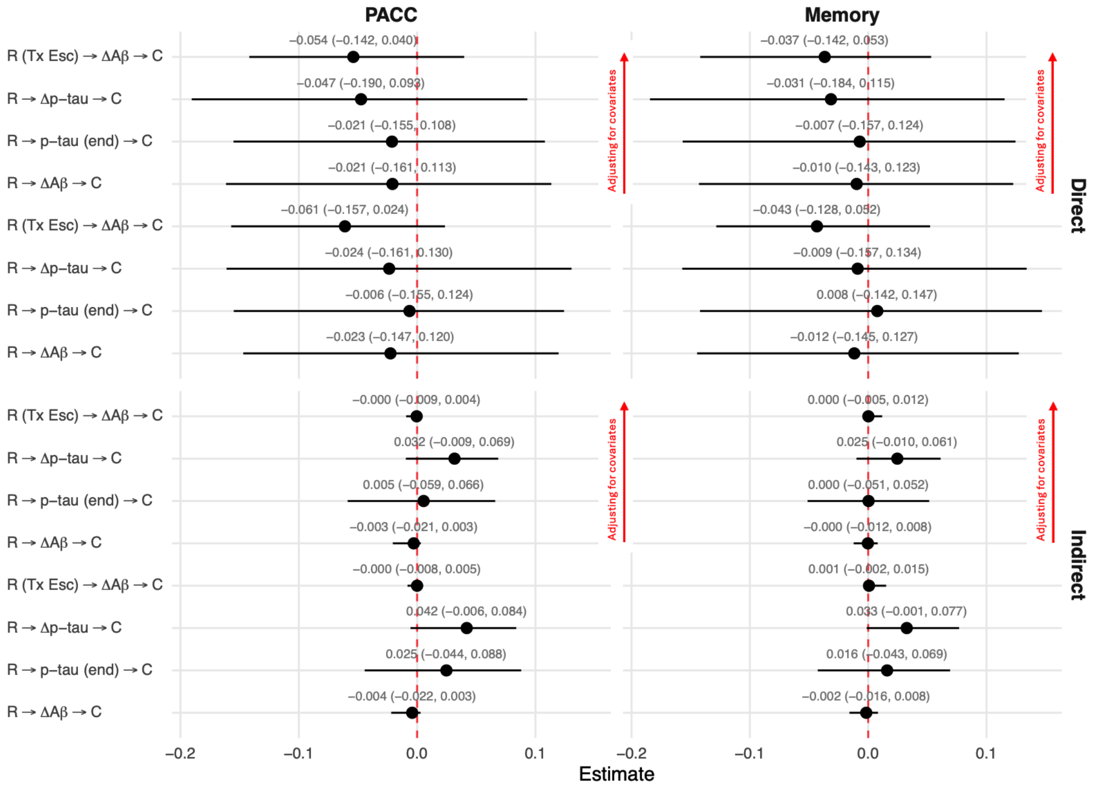


Figure S1: Estimated direct and indirect effects with 95% confidence intervals for two cognitive outcomes: PACC (left) and memory (right). Rows correspond to separate mediation models evaluating amyloid pathways. Points indicate effect estimates and horizontal bars indicate 95% confidence intervals; the dashed vertical line marks the null value. In the model labels, R corresponds to treatment randomization, R (TX esc) corresponds to pseudo-randomization, and C denotes the cognitive outcome. Mediators include change in amyloid burden ($\Delta A\beta$), end-of-treatment phosphorylated tau (p-tau end), change in phosphorylated tau ($\Delta$ p-tau).

### Supplemental References


Glymour, M. Maria (2022). "Commentary: modelling change in a causal framework". In: *International Journal of Epidemiology* 51.5, pp. 1615–1621. ISSN: 1464-3685. DOI: 10.1093/ije/dyac151.

Glymour, M. Maria et al. (2005). "When is baseline adjustment useful in analyses of change? An example with education and cognitive change". In: *American Journal of Epidemiology* 162.3, pp. 267–278. ISSN: 1476-6256, 0002-9262. DOI: 10.1093/aje/kwi187. (Visited on 11/14/2020).

Hampton, Olivia L et al. (2023). "Harmonizing the preclinical Alzheimer cognitive composite for multicohort studies." In: *Neuropsychology* 37.4, p. 436.

Mukherjee, Shubhabrata et al. (2023). "Cognitive domain harmonization and cocalibration in studies of older adults." In: *Neuropsychology* 37.4, p. 409.

Senn, Stephen (Aug. 2006). "Change from baseline and analysis of covariance revisited". In: *Statistics in Medicine* 25.24, pp. 4334–4344. ISSN: 1097-0258. DOI: 10.1002/sim.2682.

Tennant, Peter W. G. et al. (2021). *Analyses of 'change scores' do not estimate causal effects in observational data.* DOI: https://doi.org/10.1093/ije/dyab050.
